# Nano assembly of plasmonic probe-virus particles enabled rapid and ultrasensitive point-of-care SARS-CoV-2 detection

**DOI:** 10.1101/2022.08.01.22278286

**Authors:** Younggeun Park, Byunghoon Ryu, Seungjune Ki, Mingze Chen, Xiaogan Liang, Katsuo Kurabayashi

## Abstract

The current COVID-19 global pandemic caused by the severe acute respiratory syndrome coronavirus 2 (SARS-CoV-2) coronavirus has become a major public health concern. The ability to identify the virus’s presence in infected hosts with sufficient speed and sensitivity is critical to control the epidemic timely. Here, we use self-assembly of arrayed gold nanoparticles (AuNPs) on the coronavirus, which we call the “plasmo-virus particle,” to achieve a rapid, sensitive, sample preparation-free assay enabling direct detection of SARS-CoV-2 in a point-of-care (POC) setting. The AuNPs of the plasmo-virus particle serve as plasmonic nanoprobes that specifically bind to the spike protein (S-protein) sites on the surface of SARS-CoV-2. Optical interactions between the self-assembled plasmonic nanoprobes generate multiple modes of highly enhanced plasmonic coupling. Measuring changes of the multimode plasmonic coupling-induced extinction peaks allows for quantifying SARS-CoV-2 at low titers with a limit of detection (LOD) of 1.4 × 10^1^ pfu/mL. Using a miniaturized standalone biochip reading device, we further demonstrate the nano assembly assay for smartphone-operated SARS-CoV-2 detection for viral transport medium (VTM) samples within 10 min without any sample purification steps. We anticipate that the high sensitivity and speed of the POC detection performance of this biosensor technology could be broadly accepted for timely personalized diagnostics of infectious agents under low-resource settings.

## Introduction

Facing the global pandemic of acute respiratory coronavirus disease (COVID-19), healthcare workforce urgently needs the ability to detect the earliest stages of infection at the point of care (POC) or for clinical screening in high-incidence areas in a timely and cost-effective manner. The current standard COVID-19 diagnosis is based on reverse transcriptase polymerase chain reaction (RT-PCR)^1,2^. However, RT-PCR is relatively complex and expensive, making its adoption for POC testing prohibitive, where early detection of viral infection is required with little cost and time while maintaining sufficiently high levels of sensitivity and accuracy. Immunoassays are simpler, lower cost, and more rapid than RT-PCR and can be more easily deployed in an array of settings. The lateral flow test provides the most common rapid, cost-effective immunoassay platform for influenza diagnosis like a pregnancy test ^3,4^. Although several lateral flow-based POC products are commercially available, they lack the sensitivity required for detecting viral infection in early stages. Moreover, it is reported that the lateral flow test results in only 70% positive rates of RT-PCR tests ^5^. Therefore, there is a pressing need for alternative less expensive and rapid methods retaining sufficient sensitivity for COVID-19 diagnosis.

In recent years, a new class of biosensors has been demonstrated towards enabling rapid and sensitive POC COVID-19 diagnosis. COVID-19 viral detection is typically performed by analyzing either the genomic or proteomic characteristics of SARS-CoV-2, which has a viral protein-contained lipid bilayer (envelope) encapsulating a large positive-sense single-stranded ribonucleic acid (RNA) genome. Molecular testing-based COVID-19 biosensors employ viral RNA analyses coupled with photothermally induced PCR at complementary DNA-conjugated gold nano island hotspots,^6^ clustered regularly interspaced short palindromic repeats (CRISPR) machinery,^7,8^ loop-mediated isothermal amplification (LAMP),^9^ and electromechanical microcantilever detection.^10^ These analytical methods can eliminate RNA isolation/purification, temperature cycling, or even sample amplification. However, they are not as sensitive as RT-PCR or they require temperature-controlled multistep reactions, resulting in a sample processing time between 1 and 3 hours per sample at room temperature.^11-14^ Researchers have demonstrated a portable assay that enables smartphone-read CRISPR detection of amplified viral RNA with a short (15-min) assay turnaround and a low limit of detection (LOD) similar to that of RT-PCR.^15^ But such a platform still requires careful sample lysis, reagent mixing, and temperature control with a skilled user. As a result, establishing a sufficiently high level of user-friendliness for the system would be necessary to avoid user-to-user assay variation and error.

Localized surface plasmon resonance (LSPR) nanostructures have attracted a lot of interest for the highly sensitive POC biosensing method. LSPR is a strong photon-driven coherent oscillation of the surface conduction free electrons, which can be modulated when coupling occurs at the surface of the plasmonic materials.^16-18^ LSPR nano-biosensors have enabled direct detection with great sensitivity to local fluctuation, including refractive index change, charge transfer, and molecule binding, due to the amplified plasmonic field in the nanostructures. Thus, LSPR is an ideal candidate for real-time and label-free detection of micro-and nanoscale analytes.^19-25^ Moreover, the robust optical setup and detection procedures of LSPR biosensors have revealed great potential in POC diagnostic systems^26,27^. To incorporate this LSPR detection principle into POC, a systematic structural assembly ensuring the maximized plasmonic field enhancement is critical to obtain high sensitivity.^28,29^ Especially, accurate design and manufacturing of plasmonic nanoprobes targeting appropriate analytes is critical for LSPR-based biosensing of viruses and bacteria with substantially larger than the probes.

In this study, we present a nano assembly-based rapid and sensitive COVID-19 biosensor assay without sample processing. This assay allows us to directly detect the presence of SARS-CoV-2 particles and quantifies their population using a hand-held POC portable device. A one-step reagent-sample mixing of the assay spontaneously constructs an array of gold nanoparticles (AuNPs) surrounding a virus particle via self assembly. Here, antibody-conjugated AuNPs suspended in the reagent solution serve as plasmonic nanoprobes that specifically bind to S-proteins on the virus surface, forming the self-assembled AuNP-virus hybrid nanostructure, or “plasmo-virus particle.” Incident light on the plasmo-virus particle induces strong multimode plasmonic coupling with an optimized plasmonic nanoprobe size (30 nm in diameter). The multimode plasmonic coupling results in multiple LSPR peaks whose intensities vary with the virus particle population with a low LOD. The abundant presence of the plasmonic nanoprobes in the reagent solution as compared to the virus population in the sample leads to fast antibody-S-protein binding kinetics, keeping the assay time significantly short and comparable to that of a lateral flow-based POC test.

The constructed hand-held device incorporates a micro-optoelectronic unit with a biochip, a microcontroller, and data transfer unit driven by smartphone application software. The device allows us to perform user-friendly nano assembly-based testing of SARS-CoV-2 in small-volume (10 µL) viral transport medium (VTM) samples with the smartphone interface. Repeated POC testing with this device may enable continuous monitoring of the virus in the in the upper respiratory tract (nose, mouth, nasal cavity, and throat) of an asymptomatic host during an early incubation phase (**Figure 1**). The presented biosensor technology appears promising to confine the community spread of SARS-CoV-2 and treat virus carriers while fully taking advantage of the narrow time window (two to five days) before symptoms appear. It also could allow a clinical study to determine the virus population threshold between symptomatic and asymptomatic infected hosts.

**Figure 1.**
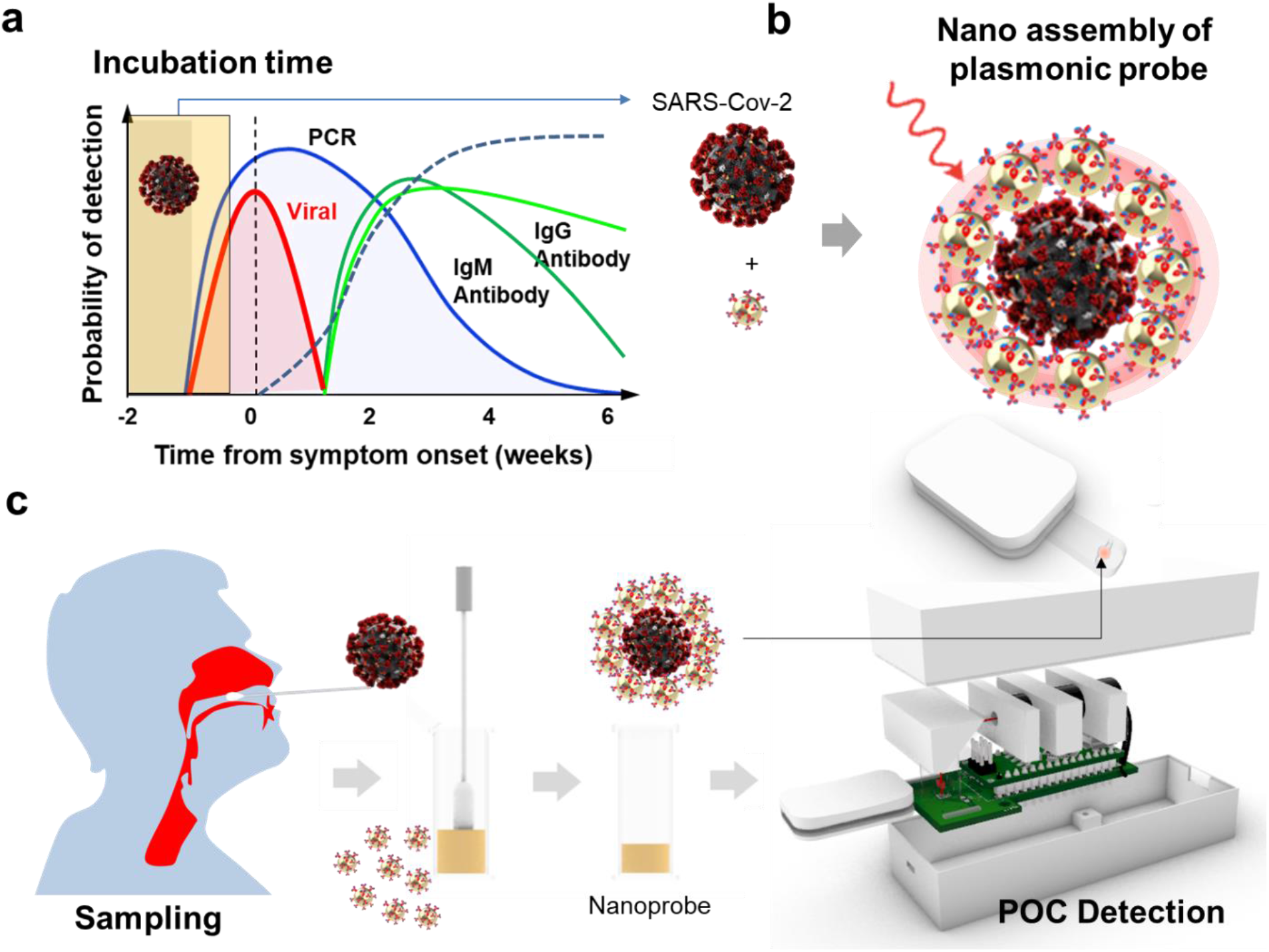
**A** point-of-care (POC) system integrated with self-assembled plasmonic virus nanostructures (“plasmo-virus particles”) for sensitive, and immediate detection of the severe acute respiratory syndrome coronavirus (SARS-CoV-2) virus. A) Timelapse vs virus detection options/target analytes; the dynamics of symptom is initiated by following exposure to the virus. B) Illustration of the plasmo-virus particle. The particle’s plasmonic nanogaps lead to a near-infrared (NIR) extinction peak. C) Detection steps with the constructed POC system and smartphone application software. The detection steps involve i) collecting a body fluid via a nasal swab, ii) mixing it with plasmonic nanoprobes before (middle left) and after (middle right) incubation, iii) loading the plasmo-virus sample into a biochip, iv) detecting optoelectronic signal with the hand-held POC system, and v) displaying the detection result on the smartphone app software.

## Results and Discussions

### Plasmo-virus particle manifests multi-mode near-field electromagnetic field (EM) enhancement

**Figure 2**. shows a schematic of the plasmo-virus particle structure. The original SARS-CoV-2 viral particle consists of a single positive-strand RNA genome encoding four structural proteins: 72 spike (S), envelope (E), matrix (M), and nucleocapsid (N). SARS-CoV-2 utilizes angiotensin-converting enzyme II (ACE2) as a cellular entry receptor during the infection process, which is also a well-known host cell receptor for SARS-CoV.^5^ SARS-CoV-2 colocalizes with ACE2 in cells.^6^ Its S-protein binds ACE2 with a high affinity.^5-9^ In addition, human antibodies bind to the viral S-protein, which prevents the virus from entering the host cell and marks it for clearance. From the geometric and functional characteristics of the infection mechanism, the S protein has been recognized as a key signature of SARS-CoV-2. Gold nanoparticles (AuNPs) conjugated with the human anti-S-protein antibodies are used as the plasmonic nanoprobes. Under such geometric arrangements, antigen-antibody binding allows the plasmonic nanoprobes to be attached onto the S-protein of SARS-CoV-2, leading to spontaneous nano assembly of the plasmo-virus particle.

**Figure 2.**
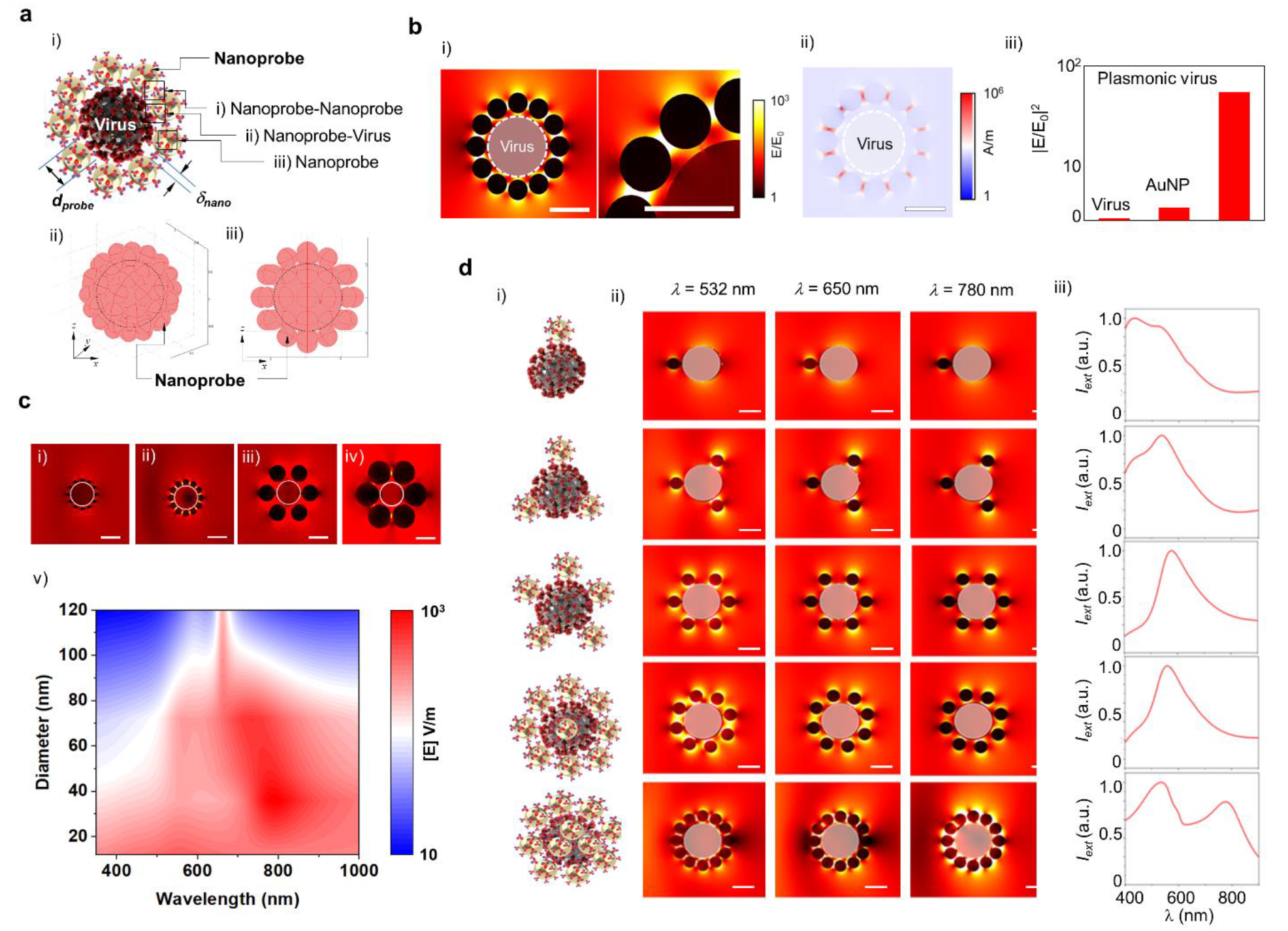
Schematic and characteristics of Plasmonic virus. **a)** Scheme of a plasmo-virus particle formed of assembled plasmonic nanoprobes with specific size and density leading to plasmonic nanogap on SARS-CoV-2 virus. **b)** i) E-field and ii) power distribution around a plasmo-virus structure. A highly enhanced E-field is focused on each plasmonic nanogap between assembled plasmonic nanoprobes on the SARS-CoV-2 virus (scale bar = 100 nm) and iii) Comparison of E-field enhancementof virus, AuNP, and plasmo-virus. **c)** E-field distribution at nanoprobes with different sizes of i) 10, ii) 50, iii) 100, and iv) 150 nm. Heat map of E-field for probe diameter vs. wavelengths from *λ* = 350 to 1,000 nm (scale bars = 100 nm). **d)** Effect of the nanoprobe density around a virus particle on the plasmo-virus particle’s optical properties; i) Schematics, ii) *E*-field distributions at 532 nm, 650 nm, 780 nm, and iii) spectra at 1, 5, 12, 32, and 72 plasmonic nanoprobes (diameter = 30 nm) on a SARS-CoV-2 virus (scale bar = 100 nm).

The nanoprobe diameter (*d*_*probe*_) and density (*n*_*probe*_) are decisive factors affecting on the plasmonic modes (**Figure 2a**). To analyze the plasmonic modes and electromagnetic (EM) field distribution in the near fields of the plasmonic nanoprobes assembled on SARS-CoV-2, we performed finite element analysis (FEA), assuming a surface of SARS-CoV-2 (*d*_*virus*_ = 100 nm) fully covered with evenly distributed nanoprobes (diameter, *d*_*probe*_ = 30 nm) (**Figure 2b**). The analysis shows multi-plasmonic modes around the plasmo-virus particle associated with three different plasmonic coupling mechanisms. First, the nanoprobe-nanoprobe interaction *via* the plasmonic nanogap (*δ*_*nano*_) (= ∼ 0.5 nm) generates strong coupling and EM field enhancement. Second, the interaction between each nanoprobe and the virus structure leads to weaker EM field. Third, the plasmonic nanoprobe itself also serves as an EM field focusing point. We found that the plasmonic coupling stemming from the nanogap effect is most influential in determining the EM field and optical properties of the assembled plasmo-virus structure.

**Figure 2c** predicts how the value of *d*_*probe*_ between 10 and 120 nm affects the EM field distribution and enhancement around the plasmo-virus particle for different incident light wavelengths (*λ* = 350 – 1,000 nm). A large EM enhancement occurs at a long wavelength of ∼ 850 nm with *d*_*probe*_ < 40 nm. A larger value of *d*_*probe*_ (≥100 nm) leads to a smaller value of *n*_*probe*_, which results in a reduction of EM field enhancement at a given wavelength in general. Additionally, we found that the predicted EM field enhancement manifests very distinct wavelength-specific behavior with an appropriate selection of the value of *d*_*probe*_. Specifically, the largest EM field enhancement (= ∼ 10^3^) occurs at *λ* between 700 and 850 nm with *d*_*probe*_ ∼ 30 nm.

With *d*_*probe*_ = 30 nm, we investigated the impact of the value of *n*_*probe*_ on the plasmo-virus particle’s EM field enhancement and optical extinction spectrum (**Figure 2d and Figure S1**). As the number of nanoprobe particles on a virus increases from 1 to 72, the inter-nanoprobe distance, *δ*_*nano*_ (Figure 2a), decreases 10 to 0.3 nm. The analysis shows that *δ*_*nano*_ = 0.3 nm yields nearly five times larger EM field enhancement than *δ*_*nano*_ = 10 nm. We also analyzed how varying *n*_*probe*_ would change the optical extinction spectrum of the plasmo-virus particle. The extinction spectrum of a virus covered with a smaller number (< 10) of nanoprobes is predicted to have a single broad peak at *λ* = 532 nm with a large full width at half maximum (FWHM) > 200 nm. As the number of the attached nanoprobes increases, a second peak grows at *λ* = 780 nm while the first peak at *λ* ∼ 550 nm becomes weaker. These trends of peak location and intensity stem from the multi-plasmonic modes described above.

### Plasmo-virus particle nano assembly enables rapid and sensitive bioassay

We experimentally characterized the nano assembly kinetics and optical property evolution of the plasmo-virus particle (**Figure 3**). First, we prepared the plasmonic nanoprobes by conjugating AuNPs with the spike protein (anti-S pro) antibody (**Figure 3a** and **see the “Experimental Section”**). We anticipated that selecting the nanoprobe size to be *d*_*probe*_ = 30 nm would maximize the nanogap plasmonic effect between the assembled nanoprobes. Our assumption here is based on the typical number of S-protein sites on a SARS-CoV-2 virus (100 nm in size) reported in the literature^30^, and it is verified by the FEA results above. The FEA results also predict a growth of the second extinction spectral peak around *λ* = 780 nm as the plasmo-virus particle assembly progresses (**Figure 3b**). The number of the nanoprobe arrays assembled on the SARS-CoV-2 virus can vary with the incubation time for a given virus population in a biofluid or with the virus population at a given incubation time.

**Figure 3.**
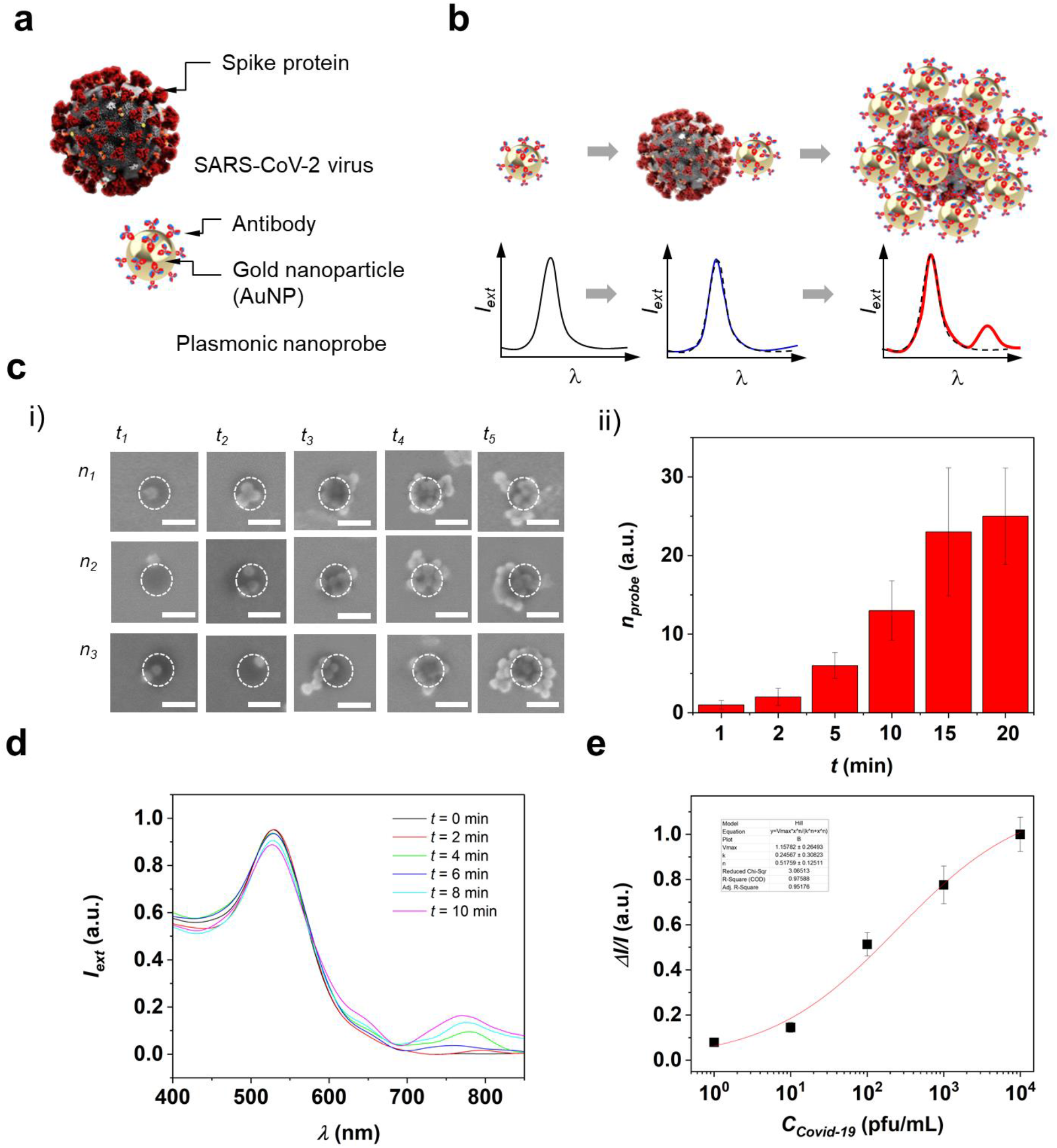
Proof of concept of plasmonic nanogap array on SARS-CoV-2 virus. **a)** Schematic of the bioconjugation of a plasmonic nanoprobe to a target S-protein on the SARS-CoV-2 virus. **b)** Spectral evolution from a single plasmonic nanoprobe to a plasmo-virus particle with an increasing number of nanoprobes attached onto the SARS-CoV-2 virus (scale bars = 100 nm). **c)** i) SEM images and ii) number of nanorpobes after different incubation times of *t*_*1*_ = 1, *t*_*2*_ = 2, *t*_*3*_ = 5, *t*_*4*_ = 10, and *t*_*5*_= 20 min (n = 5) for *C* _*SARS-CoV-2*_ = 10^3^ pfu/mL. **d)** Extinction spectra of the mixture solution of plamonic nanoprobes and SARS-CoV-2 particles after different incubation time points. **e)** Constructed calibration curve for SARs-CoV-2 detection from the spectral peak intensity at *λ* = 780 nm of figure 3c.

Subsequently, we loaded nanoprobes into a solution containing SARS-CoV-2 and took snapshot scanning electron microscopy (SEM) images of individual plasmo-virus particles at different incubation times (**Figure 3c**). The SEM images show that the number of assembled nanoprobes became saturated around the virus particles after a 20 min incubation. The rapid assembly of plasmonic nanoprobes on SARS-CoV-2 provides a biosensing approach leading to a short assay turnaround. A typical virus biosensor system employs the anti-S protein antibody as the active biological probe material immobilized on the transducer surface, and the antibody provides a surface binding site for the virus particle generating the signal. For low abundance virus in the solution, such a system requires a long assay turnaround due to slow analyte-probe kinetics accompanying the growth of a depletion region over time near the sensor surface, and the biosensor operation is governed by the analyte diffusion and convection (mass-transport-limited regime). In contrast, the plasmo-virus particle nano assembly-based assay involves surrounding the analyte virus with abundant plasmonic nanoprobes regardless of the analyte concentration. Here, the biosensing method is performed under the surface binding reaction-limited regime, where the sensor time constant *τ* is determined by the association (*k*_*on*_) and dissociation (*k*_*off*_) rates and the nanoprobes concentration (*C*_*probe*_) in the nanoprobes-virus mixture solution as *τ* = (*k*_*off*_ + *C*_*probe*_*k*_*on*_)^-1^. Thus, setting *C*_*probe*_ to make the nanoprobe population much larger than the virus population in the mixture solution allows us to keep the assay time remarkably short.

Additionally, we characterized the UV-VIS extinction spectrum for the mixture solution of plasmonic nanoprobes and SARS-CoV-2 particles at different incubation times using a spectrophotometer (Agilet 8453 G1103A Spectrometer) (**Figure 3d**). Initially, only a single peak around 530 nm was observed at *t* = 0. As the incubation process progressed, three extinction peaks started to appear at approximately *λ* = 530, 630, and 780 nm and became apparent at *t* = 10 min. These three peaks are attributed to the EM confinement in each AuNP, AuNP -SARS-CoV-2 plasmonic coupling, and AuNP-AuNP plasmonic coupling, respectively. The acquired spectra matched well the theoretical predictions in **Figure 2**. We further analyzed the intensity variation of the extinction peak at *λ* = 780 nm for the spectrum at *t* = 10 min with the number of the virus particles (*C*_*SARS-CoV-2*_) in the solution varying from 10^0^ to 10^4^ pfu/mL and constructed a standard calibration curve (**Figure 3e**). From this curve, the LOD value was estimated as LOD = 3*σ*/*k*_*slope*_, where *σ* and *k*_*slope*_ are the standard deviation of the background signal measured from a blank control and the regression slope of the calibration curve, respectively. We found that the assay achieved a LOD of 7.6 × 10^1^ pfu/mL with a large (10^4^) dynamic range.

### Plasmo-virus particle nano assembly-based bioassay enables POC virus detection

We built a hand-held biosensing system for SARS-CoV-2 virus detection based on the plasmo-virus particle nano assembly assay (**Figure 4)**. The system allows us to measure the variation of light transmission through a sample solution with a complementary metal oxide semiconductor (CMOS) photodetector (**Figure 4a**). The NIR extinction of the plasmo-virus particle overlaps with the spectral responsivity of the employed CMOS photodetector (**Figure S1**). Without SARS-CoV-2 particles, the incident light at *λ* = 780 nm penetrates the on-chip solution with the highest transmission and reaches the underlying photodetector to the maximum extent. If SARS-CoV-2 particles are present, plasmo-virus particles are formed in the sample solution. Strong plasmonic coupling between the assembled plasmo-virus particles and the incident light results in an increase in the mixture solution’s absorbance. The CMOS photodetector’s photocurrent signal change is correlated with *C*_*SARS-CoV-2*_ prior to loading plasmonic nanoprobes. The quantification of SARS-Cov-2 particles with the biosensing system requires no sample preparation other than nanoprobes-sample mixing and no sophisticated optics, which makes the system desirable for POC operations.

**Figure 4.**
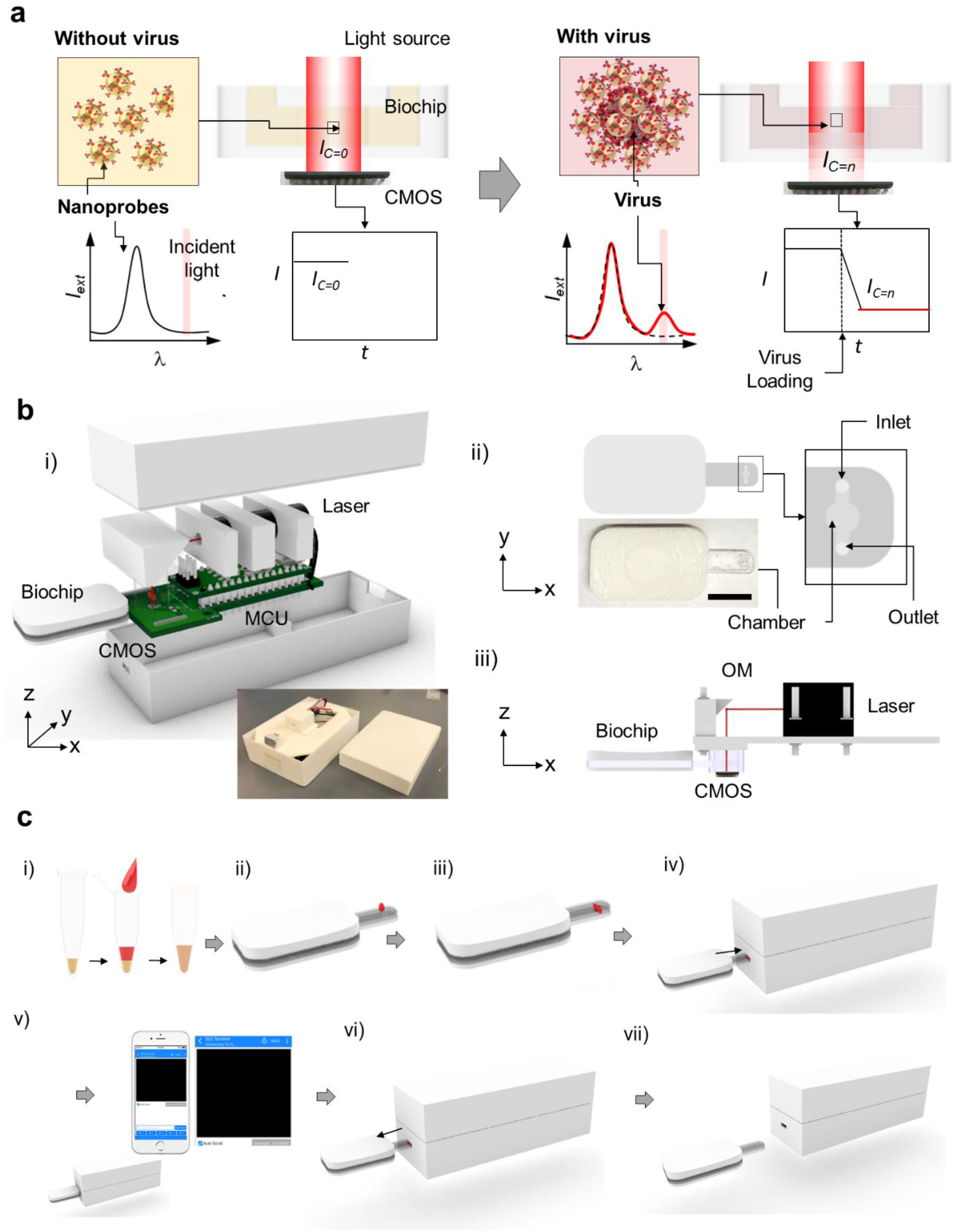
Handheld POC biosensor incorporating the plasmo-virus particle-based assay. **a)** Principle of plasmo-virus particle-based colorimetry using the handheld reading device. The sample solution suspending plasmonic nanoprobes without the virus is transparent to the incident light at *λ* = 780 nm, resulting in a higher CMOS photosensor signal (left). The nano assembly of plasmo-virus particles causes the sample to increase the light absorption, resulting in a reduction of the CMOS photosensor signal (right). **b)** i) Integrated POC biosensor module for colorimetric SARS-CoV-2 detection. The system consists of a biochip, light source, CMOS photodetector, microcontroller unit (MCU), and Bluetooth module with an integrated circuit board. ii) Diagnostic biochip cartridge loaded with a mixture solution of plasmonic nanoprobes and SARS-CoV-2 in VTM. iii) Optical arrangement with alignment between the upper light source, biochip, and CMOS detector. Light from a laser diode illuminates the biochip via an OM (optical mirror). **c)** Biosensor operation procedures; i) mixing a virus-spiked VTM solution with a plasmonic nanoprobe reagent in a PCR tube, ii) loading the mixture to the biochip cartridge, iii) incubating the mixture for 10 min, iv) inserting the biochip into the integrated POC biosensor system, v) detecting the CMOS photosensor signal from the sample and wirelessly communicating data with a smartphone, vi) removing the biochip chartridge, and vii) preparing the next measurement. Custom-made application software (Blynk Internet on thing (IoT) Platform) is used for acquiring real-time data and displaying the analyzed results.

The system (L×H×W = 10×4×2 cm^3^) consists of a light source (laser, wavelength = 780 nm and P = 1 mW), a biochip (volume_max_ = 75 µL), a lithium-ion battery (V = 3.2 V and P = 1.48 Wh), and an integrated printed circuit board that embedded a CMOS photodetector, microprocessor, and Bluetooth module (**Figure 4b, Figure S2-S4**). The light source was selected to match the extinction wavelengths of the assembled plasmo-virus particles. This monochromic light enables us to enhance the detection sensitivity due to the decreased environmental noise. The micro-optics of the system permits precise light alignment between the biochip, light source, and photodetector, and it accommodates sensitive detection of a small volume sample loaded on the biochip (**Figure S5, S6**). In addition, the microcontroller unit and the Bluetooth module enables the remote wireless operation of the constructed POC biosensing system.

**Figure 4c** shows the on-chip assay process using the system. Prior to implementing this assay, we optimized the geometry of the biochip and the concentration of the nanoprobe solution to achieve a optimal optical response to ON/OFF switched incident light on a portable scale (**Figure S7**). The process started with mixing a plasmonic nanoprobe reagent and a virus-spiked VTM sample solution in a 0.2 mL PCR tube. The nanoprobe-sample mixture was loaded onto the biochip, and the biochip was inserted into the entrance of the integrated POC biosensing system, followed by detection of the photocurrent signal. We used custom-made software for smartphone-based real-time signal monitoring and operation control of the system.

### Rapid POC SARS-CoV-2 detection achieves ultrahigh sensitivity

Finally, we characterized the SARS-CoV-2 particle detection performance of the constructed biosensing system (**Figure 5**). After loading the mixture solution of plasmonic nanoprobes and SARS-CoV-2 particles on the biochip, we inserted it into the POC system and measured the temporal photoresponse under light illumination (*λ* = 780 nm) at different values of *C*_*SARS-CoV-2*_ ranging from 10^0^ to 10^4^ pfu/mL. For the entire range of *C*_*SARS-CoV-2*_ tested, the normalized photocurrent signal change (*I*_*norm*_ = Δ*I/I*_*0*_) reached a plateau within 10 min (**Figure 5a**). Subsequently, we obtained a calibration curve based on the plasmonic mode at *λ* = 780 nm with a 10 min incubation (**Figure 5b**). This single-mode calibration curve shows highly consistent detection characteristics for the *C*_*SARS-CoV-2*_ range with a LOD of 1.37×10^1^ pfu/mL. Recall that the plasmo-virus particle has multi plasmonic modes at *λ*=532 and 780 nm. We characterized *I*_*norm*_ at these two wavelengths and obtained a new standard calibration curve for improved detection sensitivity (**Figure S8**). The slope (*k*_*multi*_) of the multimode standard calibration curve is ∼ 2.5 times steeper than that from the single plasmonic mode curve, while the noise level (*σ*_*multi*_) increases by a factor of ∼1.5. The LOD estimated for the multi-plasmonic mode results in *C*_*SARS-CoV-2*_ = 0.5 × 10^1^ pfu/mL that is five times lower than that from the single plasmonic mode measurement. We detected SARS-Cov-2 particles in PBS, saliva, VTM/Saliva (v/v=50%), and VTM and compared the LODs for these media. The LODs are between 0.7 ×10^0^ to 1.5×10^1^ pfu/mL, which are lower than the LOD of a field-effect transistor (FET) biosensor achieving sensitivity comparable to that of LAMP-based COVID-19 diagnosis^31^ and seven orders magnitude lower than the peak number of the virus in the throat saliva of COVID-19 patients (1-2 × 10^8^ pfu/mL).^32^ It should be noted that VTM exhibits an extinction peak at 550 nm (**Figure S9**), and the strong extinction peak of nanoprobes is distinguishable. The NIR colorimetric detection yields consistent detection performance regardless of the mediuam type. Furthermore, to test the specificity of our POC SARS-Cov-2 biosensor, we measured the sensor signal (*I*_*norm*_) for VTM samples containing microphase (MS_2_), cowpea mosaic virus (CPMV), SiO_2_ nanoparticles (SiNP), or SARS-Cov-2 at different populations ranging from 10^0^ to 10^4^ particles/mL (**Figure 5d**). We observed negligible signals from the non-SARS-CoV-2 samples as compared to the signals from SARS-CoV-2 samples (P = 0.001). This verifies the negligible cross-reactivity of the SARS-CoV-2 targeting plasmonic nanoprobes.

**Figure 5.**
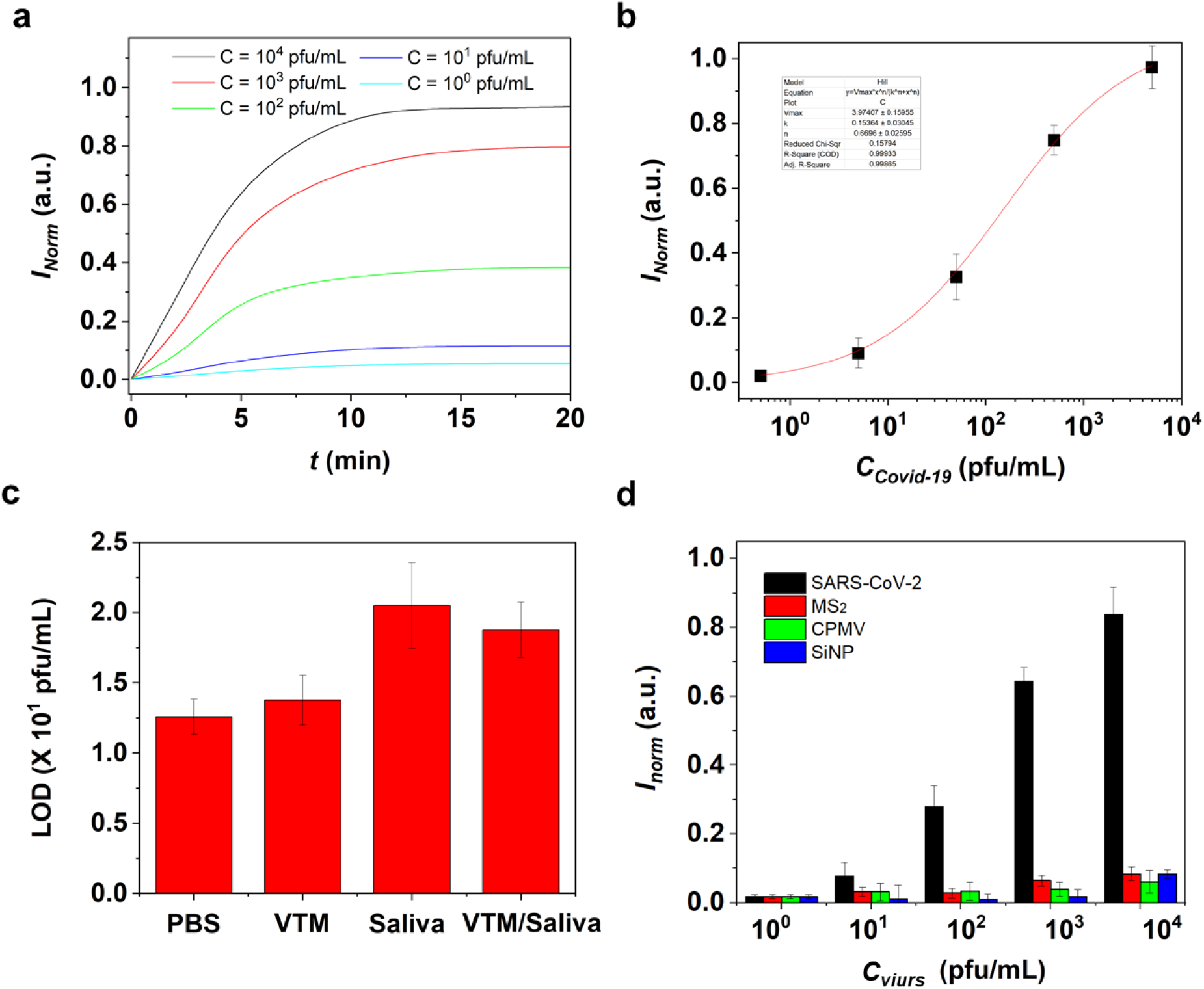
Detection performance of the integrated biosensor system for virtual transfer media (VTM) samples. **a**) Temporal responses of the normalized biosensor signal (*I*_*norm*_ = *ΔI/I*_*0*_) that represents the nanoprobe-virus binding kinetics for samples spiked with different virus concentrations (unit of virus concentration: pfu/mL). **b**) Calibration curve of C_SAR-Cov-2_ (n = 5) obtained by the biosensor system with NIR incident light (*λ* = 780 nm). **c**) Biosensor LODs for PBS, Saliva, VTM/Saliva (v/v=50%), and VTM. **d**) Normalized biosensor signals (*I*_*norm*_) for VTM solutions, each spiked with one of four analytes: SARS-CoV-2, MS_2_, CPMV, and SiO_2_ nanparticles (diameter = 50 nm) at various concentrations. The particle concentration unit for SiO_2_ is particles/mL. Error bars represent the standard deviation errors (n = 5, P < 0.05).

### Conclusion

We successfully demonstrated a AuNP-virus nano assembly-based assay that enables sensitive and rapid detection of SARS-CoV-2. The assay protocol only involved one-step reagent-sample mixing and subsequent optoelectronic detection, thus eliminating laborious procedures which are typical of PCR test, including sample lysis, RNA isolation, and multistep enzymatic reactions. The plasmonic nanoprobes assembled on the SARS-CoV-2 spike protein sites formed multidimensional nanogap arrays on the virus. These plasmonic nanogap arrays resulted in highly enhanced plasmonic modes. The combination of the multi-plasmonic modes allowed the sensitive detection of SARS-CoV-2 as low as LOD = 1.4 × 10^1^ pfu/mL. The snapshot SEM images taken after different incubation times revealed the rapid kinetics of the plasmonic nanoprobe self-assembly on the virus surface, which allowed the virus particle detection to be completed within 10 min. In addition, we incorporated the nano assembly-based assay into a hand-held portable detector device built with a micro-optics unit. The hand-held device sustained the highly sensitive detection performance of SARS-CoV-2 with a small volume of 10 uL. Furthermore, the hand-held device permitted real-time assay signal monitoring in the presence of SARS-CoV-2 in clinically relevant media via smartphone-based wireless data transmission and analysis. The speed, sensitivity, portability, and user-friendliness of the demonstrated assay may find wide use in POC detection of various pathogenic organisms and infectious agents besides SARS-CoV-2.

## Method

### Synthesis of plasmonic nanoprobe

We functionalized the thiolated alkane 10-Carboxy-1-decanethiol (HS-(CH2)10-COOH) with AuNPs (0.2 nM). Then, using standard 1-ethyl-3-[3-dimethylaminopropyl] carbodiimide / N-hydroxysuccinimide (EDC/NHS) coupling chemistry, we performed bioconjugation to bind antibody on the AuNP surface. To prevent non-specific binding from fouling the AuNP surface, we treated the produced antibody-AuNP conjugates with 1% BSA in 1x PBS in blocking solution and incubated the entire system for 20 minutes. Using 20L of 1PBS, the antibody-AuNP particles were carefully washed three times to eliminate any surplus solutions or molecules.

### Numerical anlaysis of electromagnetic field

By solving the Helmholtz wave equation, we performed a finite element analysis (FEA) to evaluate the near-field electromagnetic fields surrounding a scattered AuNP and assembled AuNPs on a SARS-CoV-2 virus (see **Supporting information**).

### Construction of POC diagnosis system

A micro-optics, system control, and wireless communication module comprise the portable POC diagnostic system.

#### Micro-optics module

We built a fully integrated micro-optics module that includes a biochip, a light source (Lucky light, LL-S150W-W2-1C, I = 350 mcd), a commercial CMOS photodetector (ams, TSL2591), a mirror, and a micro-optical cage (see **Supporting information**).

#### System control module

We designed and prepared a printed circuit board (PCB) (W×L×H = 64.7 × 31.2 ×1 mm^3^) to embed an Arduino Nano (WYPH, Arduino Nano) with a microcontroller (MCU, ATMEGA328P), a Bluetooth BLE (DSD Tech, HM-10), and Li-ion battery (2500mA and 3.7 V) (see **Supporting Information**).

#### Wireless communication and application software module

We used an application software incorporated in a smartphone (Apple, iPhone 8) for remote operation of the POC diagnostic system and real-time data presentation. The smartphone app communicates with the system through Bluetooth BLE in the microcontroller settings.

### Sample loading and viral particle analysis

For on-chip tests using VTM clinical samples and spiked VTM samples, the sample was loaded into a 100 µL microtube. The identical volume of the plasmonic nanoprobe reagent was made off-chip and then put into the microtube containing the sample. After 10 minutes of incubation, the mixed sample was put onto the chip. Once the sample and reaction chemicals were fed into the chip, it was placed in the POC diagnostic system’s micro-optics module. We detected the fed sample by running the POC system. The application program then displayed the infection level in 1 second.

## Data Availability

All data produced in the present study are available upon reasonable request to the authors

## Supporting Information

This file contains supporting methods and figures.

## Acknowledgements

This work was supported by the National Science Foundation, NSF-CBET program (2030551) and Academic Research Fund and Mcubed (U064088) at the University of Michigan.

