## Supporting information for "Nano assembly of plasmonic probe-virus particles enabled rapid and ultrasensitive point-of-care SARS-CoV-2 detection"

Dr. Younggeun Park

Department of Mechanical Engineering

Weil Institute for Integrative Research in Critical Care

University of Michigan, Ann Arbor, MI 48109

Prof. Xiaogan Liang

Department of Mechanical Engineering

Weil Institute for Integrative Research in Critical Care

University of Michigan, Ann Arbor, MI 48109

Prof. Katsuo Kurabayashi

Department of Mechanical Engineering

Weil Institute for Integrative Research in Critical Care

Department of Electrical Engineering and Computer Science

University of Michigan, Ann Arbor, MI 48109

### List of contents

**Methods**

**Figure S1. CAD design of the integrated POC biosensor system.**

**Figure S2. Integrated circuit block diagram.**

**Figure S3. Integrated electric circuit board;** a) circuit design and b) integrated board to embed a CMOS detector, a micro control unit (MCU), a Bluetooth module, and a powder module.

**Figure S4. Photo images of the integrated POC biosensor system;** a) integrated circuit in the opened packaging and b) integrated microoptic module.

**Figure S5. System-level ON/OFF optical signal test using the integrated biosensor system.**

**Figure S6. Impact of the biochip chamber height on the biosensor detection sensitivity.** The plots represent calibration curves for the standard agent (methylene blue) ranging from 0.01 to 100 mMat with the biochip chamber height *h* of 1, 2, and 3 mm.

**Figure S7. Impact of the plasmonic nanoprobe density on the biosensor signal for SARS-CoV-2.** The data show the normalized CMOS phososensor signal values obtained for the SARS-CoV-2 population of 0.001, 0.1, and 10 PFU/μL with the plasmonic nanoprobe optical density (OD) of 0.1, 0.5, 1, and 5.

**Figure S8. Calibration curves of the integrated biosensor system for SARS-CoV-2 obtained by visible (VIS) incident light (Black) and near infrared-infrared (NIR-I) incident light (Red).**

**Figure S9. Extinction spectrum of VTM.** There is no spectral overlap between the plasmo-virus particle and VTM.

**Methods**

**Chemical and bioagents:** We purchased gold nanoparticle (AuNPs, *d* = 40 nm) from Tedpella. 10-Carboxy-1-decanethiol (C-10), and albumin, from bovine serum (BSA), were purchased from Sigma Aldrich. 1-ethyl-3-[3-dimethylaminopropyl] carbodiimide (EDC) and/ N-hydroxysuccinimide (NHS) were purchased from ThermoFirscher Co. For experiments using the virus, heat-inactivated SARS-related coronavirus 2, CPMV, and $MS_2$ were purchased from ATCC. SiNPs were synthesied by Stöber method. SARS-CoV-2 spike antigen protein (40591-V08H) and SARS-CoV-2 spike antibody (40150-R007) were purchased from Sino Biological, Inc., China. respectively. Nano pure deionized (DI) water (18.1 MΩ-cm) was produced in-house.

**Synthesis of antibody-conjugated AuNP plasmonic nanoprobes:** In the beginning, we centrifuged a solution suspending gold nanospherical particles (AuNPs) (0.2 nM) three times at 5,000 rpm for 10 min and washed the AuNPs in D.I. water to remove excessive structure direction agents (citrate) from the solution. Subsequently, we functionalized the AuNPs with thiolated alkane 10-Carboxy-1-decanethiol (HS-($CH_2$)10-COOH) using a self-assembly method (SAM). In this process, AuNP colloidal solution was first incubated in 1mM of thiolated alkane 10-Carboxy-1-decanethiol (HS-$(CH_2)_{10}$-COOH) overnight. Then, the antibody was linked to the –COOH functional group formed on the AuNP surface by means of standard 1-ethyl-3-[3-dimethylaminopropyl] carbodiimide / N-hydroxysuccinimide (EDC/NHS) coupling chemistry. Here, we first washed the –COOH functionalized AuNPs and loaded these AuNPs into a mixture of 0.4 M EDC and 0.1 M NHS at a 1:1 volume ratio in a 0.1 M EDC solution to activate the AuNP surface. We mixed a solution of primary S-protein antibodies diluted from 100 to 10 μg/mL in 1x PBS with the solution of the AuNPs functionalized above in a PCR tube and incubated it for 60 min. Subsequently, we treated the prepared antibody-AuNP conjugates with 1% BSA in 1x PBS in a blocking buffer. Then, we incubated the whole solution for 20 min to suppress AuNP surface fouling due to non-specific binding. Before measuring SARS-Cov-2 virus, these antibody-AuNP nanoprobes were thoroughly washed three times using 20μL of 1×PBS to remove any excessive solutions or molecules. In addition, we obtained the extinction spectrum of the AuNP colloidal solution before and after the functionalization using a UV-VIS spectrometer (Agilent 8453 G1103A Spectrophotometer) to confirm that the AuNPs were surely biofunctionalized.

**Electromagnetic field simulation:** We performed a finite element analysis (FEA, COMSOL Multiphysics software) to predict the near-field electromagnetic fields around a dispersed AuNP and assembled AuNPs on the SARS-CoV-2 virus by solving Helmholtz wave equation. According to the SEM images in Figure 2, we estimated the dimensions of the assembled structure. We constructed hybrid mesh structures for the AuNPs and SARS-CoV-2 virus to fit their round shapes. We tested different mesh structures for the varying number of AuNPs around SARS-CoV-2. In this analysis, we assumed the relative permeability and complex permittivity of gold to be 1 and $\varepsilon_r = f(\lambda)$, respectively. In addition, the model assumed perfect absorption at the outer boundary to minimize spurious reflections, thereby setting a perfectly matched layer and an integration layer in a concentric space.

**Construction of POC diagnosis system:** A micro-optics, system control, and wireless communication module comprise the portable POC diagnostic system.

*Micro-optics module:* A biochip, a light source (Lucky light, LL-S150W-W2-1C, I = 350 mcd), a commercial CMOS photodetector (ams, TSL2591), a mirror, and a micro-optical cage were all constructed into a completely integrated micro-optics module. The poly(methyl methacrylate) (PMMA) biochip has an input, an optical window, and an outlet. The top surface of the optical winodow was built flat to reduce light refraction from the solution surface. The micro-optical cage made of polylactic acid (Overture PLA filament 1.75mm) and printed on a 3D printer (Prusa research, Prusa I3 MK3S) serves to orient the light source, detection chamber, and photodetector with minimal dispersion.

*System control module:* We developed a PCB (WLH = 64.7 31.2 1 mm3) to accommodate an Arduino Nano (WYPH, Arduino Nano) with a microcontroller (MCU, ATMEGA328P), Bluetooth BLE (DSD Tech, HM-10), and Li-ion battery (2500mA and 3.7 V) (see Supporting Information). The inter-integrated circuit (I2C) protocol is used to connect with the Arduino Nano. The photodetector and logic voltage are powered by 3.3 V and 5 V operation voltages, respectively. We encased the integrated system in two packaging boxes (up and down) made with a 3D printer (Prusa research, Prusa I3 MK3S).

*Wireless communication and application software module:*We used an application software incorporated in a smartphone (Apple, iPhone 8) for remote operation of the POC diagnostic system and real-time data presentation. The smartphone app communicates with the system through Bluetooth BLE in the microcontroller settings.

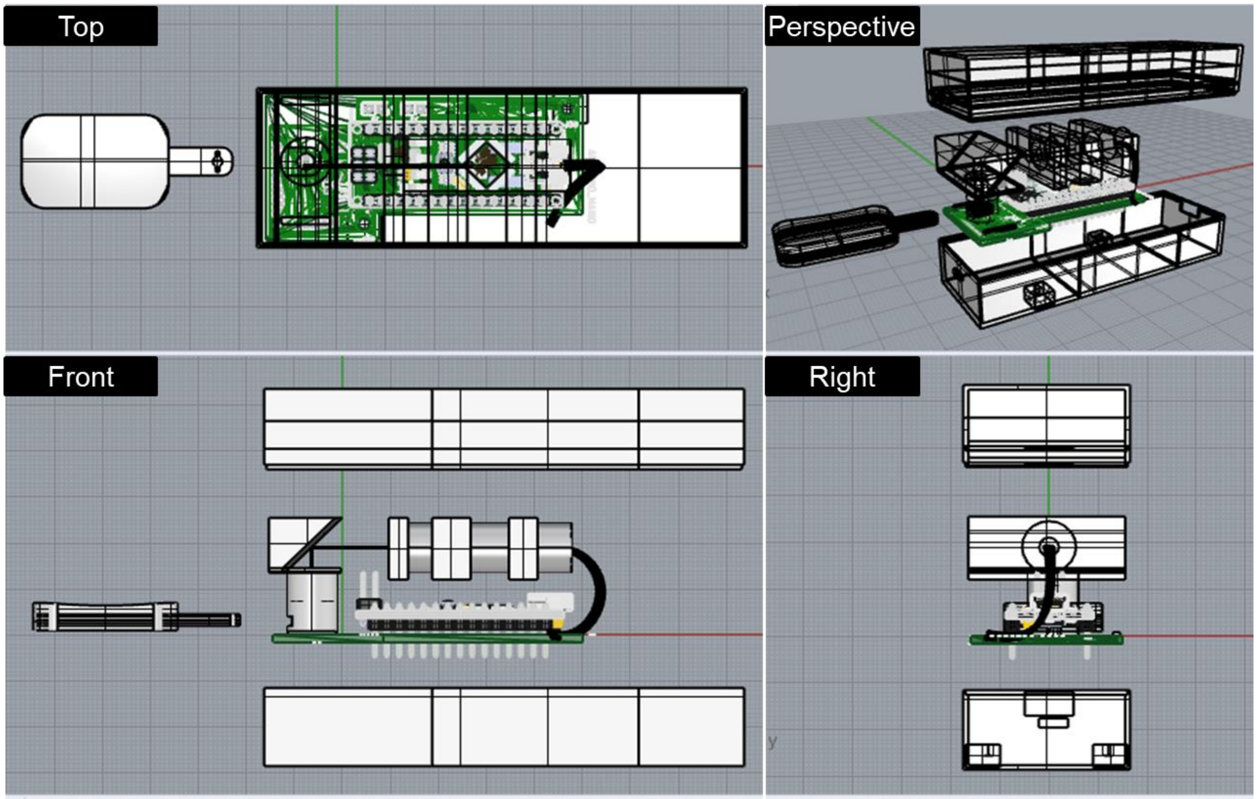


**Figure S1. CAD design of the integrated POC biosensor system.**

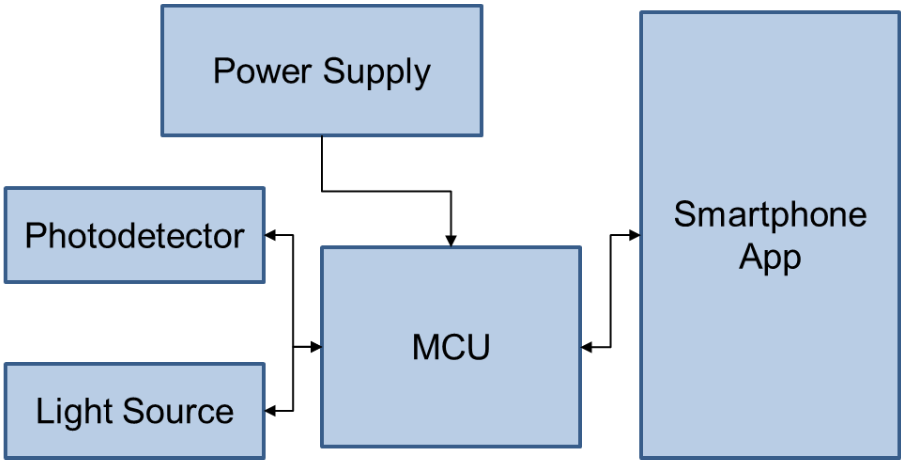


**Figure S2. Integrated circuit block diagram.**

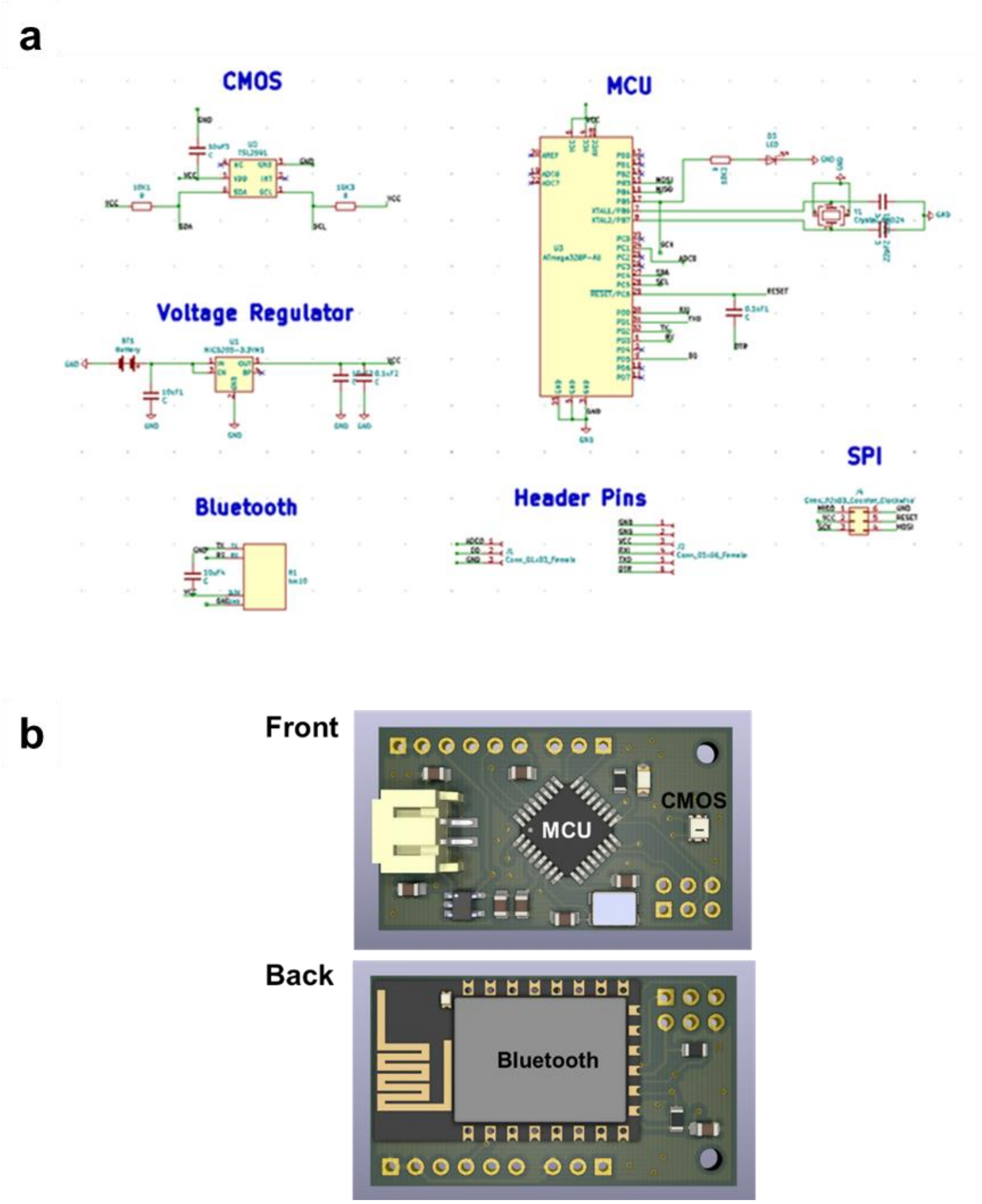


**Figure S3. Integrated electric circuit board;** a) circuit design and b) integrated board to embed a CMOS detector, a micro control unit (MCU), a Bluetooth module, and a powder module.

**a**

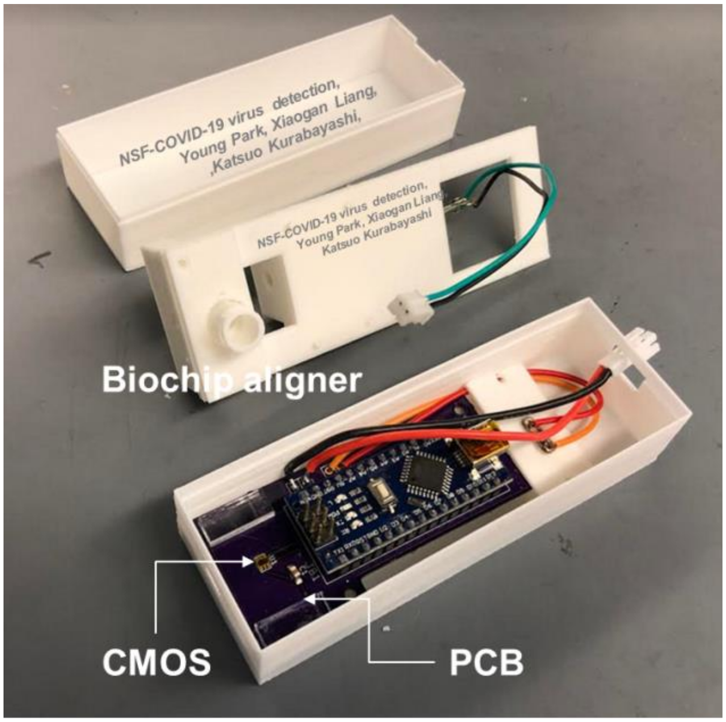


**b**

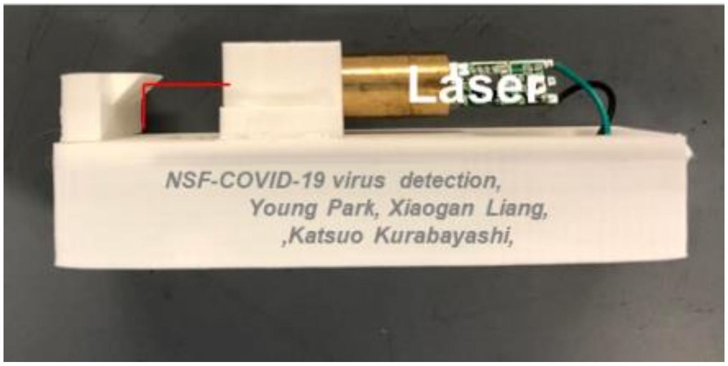


**Figure S4. Photo images of the integrated POC biosensor system;** a) integrated circuit in the opened packaging and b) integrated microoptic module.

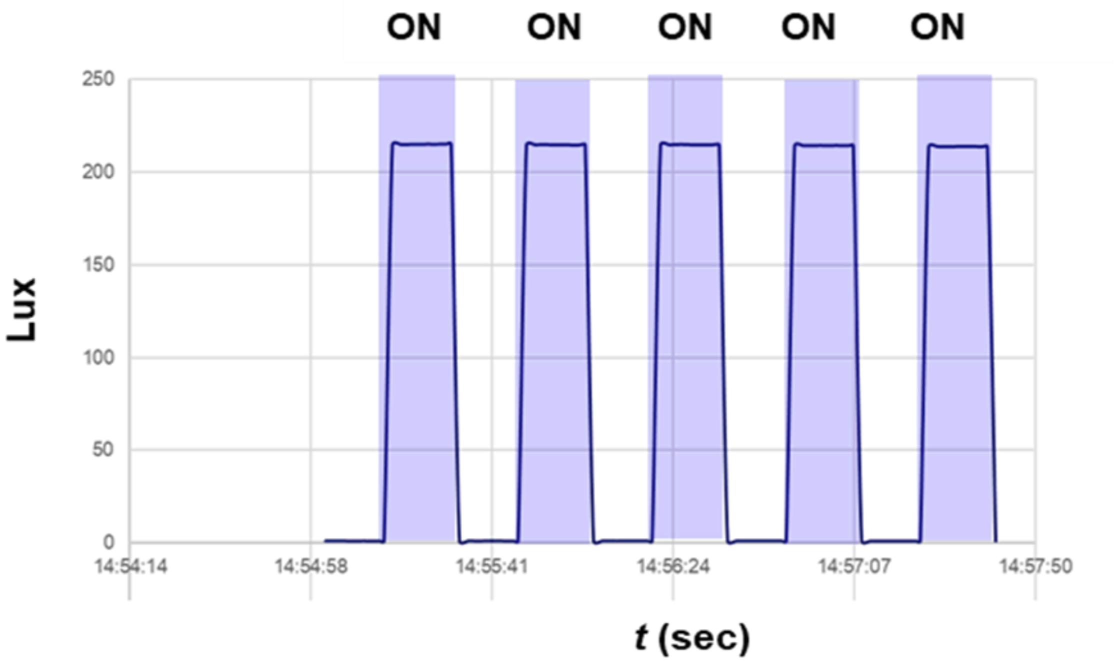


**Figure S5. System-level ON/OFF optical signal test using the integrated biosensor system.** The COMS photosensor of the integrated biosensor system effectively detected the light intensity alternating between the ON/OFF states.

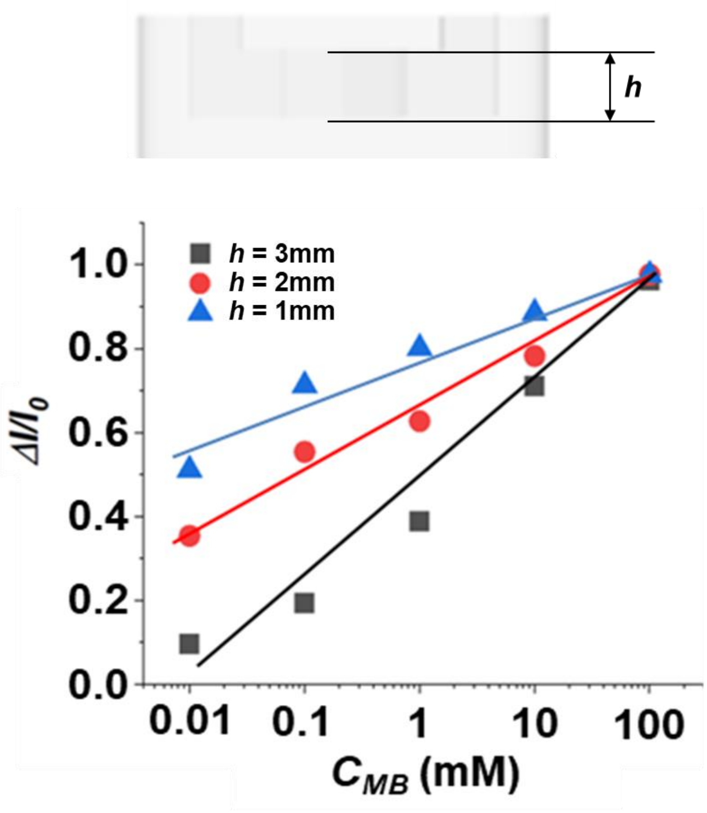


**Figure S6. Impact of the biochip chamber height on the biosensor detection sensitivity.** The plots represent calibration curves for the standard agent (methylene blue) ranging from 0.01 to 100 mMat with the biochip chamber height *h* of 1, 2, and 3 mm.

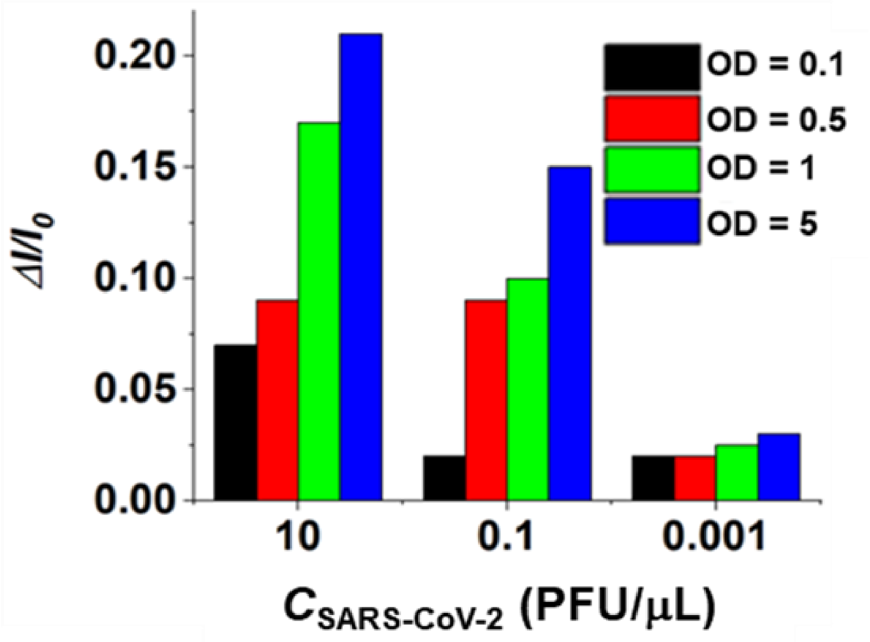


**Figure S7. Impact of the plasmonic nanoprobe density on the biosensor signal in SARS-CoV-2 detection.** The data show the normalized CMOS phososensor signal values obtained for the SARS-CoV-2 population of 0.001, 0.1, and 10 PFU/μL with the plasmonic nanoprobe optical density (OD) of 0.1, 0.5, 1, and 5.

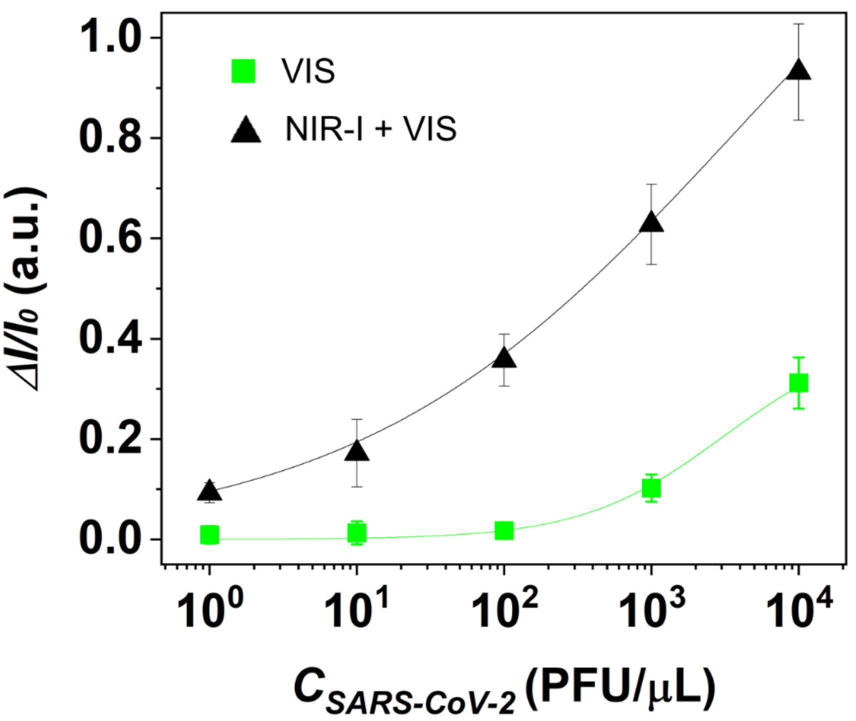


**Figure S8. Calibration curves of the integrated biosensor system for SARS-CoV-2 obtained by visible (VIS) incident light and combined near infrared-infrared (NIR-I) and VIS incident light.**

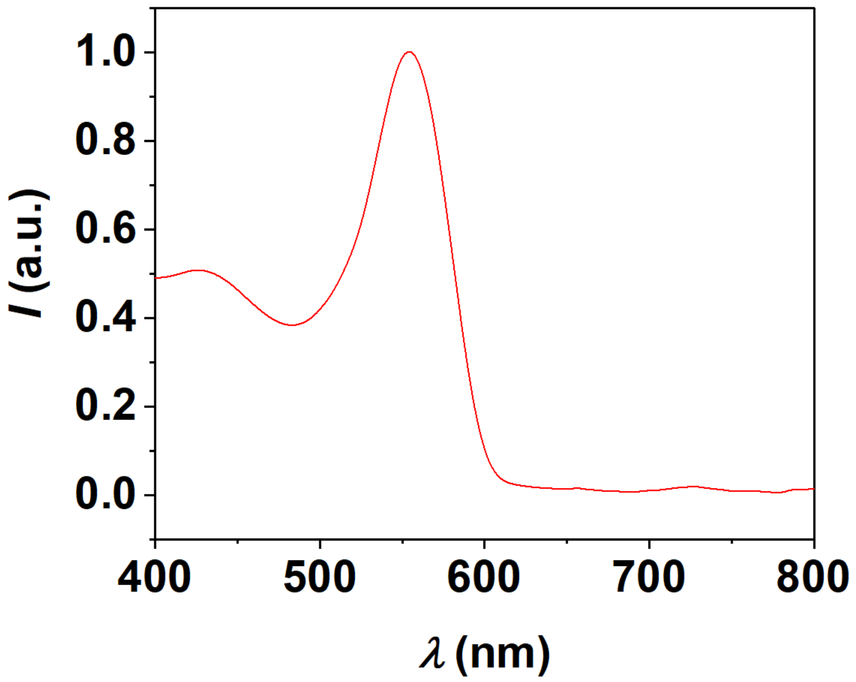


**Figure S9. Extinction spectrum of VTM.** There is no spectral overlap between the plasmo-virus particle and VTM.
